# Co-administration of prism adaptation and methylphenidate needs striatal integrity to alleviate spatial neglect

**DOI:** 10.1101/2023.01.12.23284487

**Authors:** Maude Beaudoin-Gobert, Faustine Benistant, Maxence De Lanversin, Jules Javouhay, Sophie Jacquin-Courtois, Gilles Rode, Yves Rossetti, Jacques Luauté

## Abstract

**Context:** A previous study demonstrated a long-term functional improvement of spatial neglect after methylphenidate combined with prismatic adaptation in a group of patients suffering from left spatial neglect after a right stroke (RITAPRISM study).

**Objective:** we hypothesized that the functional improvement after MP combined with PA depends on striatal integrity in responders patients.

**Methods:** We conducted an MRI study in “MP+PA” program to identify lesional pattern in responders and non-responders patients in the RITAPRISM cohort. Using anatomical segmentation on morphological MRI, we compared lesional pattern in the striatum between responders and non-responders patients.

**Results:** The beneficial effect of MP+PA co-administration should require striatal integrity in neglect patients. More specifically, our results suggest that the short-term effect is mediated by the ventral striatum whereas the long-term effect is mediated by the posterior putamen.

**Conclusion:** Benefical effet of MP+PA could rely on reinforcement processes at early stage of the MP+PA program and visuospatial substrates at long-term.

## 1 Introduction

Spatial neglect is defined as a difficulty in detecting, responding or directing attention to stimuli presented or represented on the side contralateral to a brain lesion, particularly the right hemisphere. It is a factor of poor functional evolution after a stroke (for review Luaute et al. 2006). The gold standard for the medical management of these patients is prismatic adaptation (Rossetti et al. 1998). However, a double-blind trial shows that a weekly session for four weeks is insufficient to produce a sensorimotor benefit (Rode et al. 2015). It is therefore important to define how to improve the effectiveness of prismatic adaptation in rehabilitation. In particular, efforts should focus on the need to target spatial and non-spatial attention deficits to reduce the symptoms of spatial neglect.

As methylphenidate (MP) improves non-spatial attention in patients with acquired brain injury (Kim et al. 2006), it appears as a relevant candidate to improve spatial neglect.

A previous study demonstrated a long-term functional improvement after MP combined with PA in a group of patients suffering from left spatial neglect after a right stroke (Luauté et al. 2018). However, there was heterogeneity in the clinical improvement after prismatic adaptation in patients within the “MP+PA” group with the identification of responders and non-responders. We can wonder in what extent the lesional pattern within some keys brain structures can explain this heterogeneity and lesion location is thus probably a crucial factor influencing the efficiency of the catecholamine (Luaute et al. 2006).

Among these key structures, the striatum seems particularly promising as it contains the highest density of dopaminergic transporters which MP acts on to increase the dopamine level (N. D. Volkow et al. 2002). MP blocks dopamine uptake and enhances dopamine signaling in the cortico-striatal circuit. More specifically, Positon Emission Tomography (PET) imaging in healthy subjects and ADHD patients has revealed that MP increases the synaptic concentration of dopamine mostly inside the anterior striatum (Nora D. Volkow et al. 2012; Kodama et al. 2017; Hong et al. 2015).

Thus, we hypothesized that the functional improvement after MP combined with PA depends on striatal integrity in responders patients. To do so, we conducted an MRI study in “MP+PA” program to identify lesional pattern in responders and non-responders patients.

## 2 Material and methods

### 2.1 Participants

This study was conducted in the RITAPRISM cohort (Luauté et al. 2018). Briefly, patients included in the RITAPRISM cohort fulfilled the following criteria: age above 18 and less than 80 years-old, right-handed, ischemic or haemorrhagic right stroke (>1 month, <18 months after the incident); as previously described in Luauté et al (2018) spatial neglect was confirmed by classical paper and pencil tests (line bisection, line cancellation, balloon-test, copy of a scene, drawing a daisy from memory).

### 2.2 RITAPRISM study design

The RITAPRISM study was a multicentre, double-blind and placebo-controlled study (Luauté et al. 2018). All patients provided written informed consent. The study was approved by Lyon Ethics Committee (CPP Sud-Est IV; EudraCT Number: 2008-000325-00). After inclusion, patients were randomized into a “placebo” or a “methylphenidate” (MP) group (respectively, n=6 and n=10; Figure 1). Spatial neglect was assessed in the pre-test session (D7) and during the placebo/methylphenidate treatment in post-tests sessions at the early (D15), intermediate (D22) and final (D45) phase. Patients performed the line bisection measure. The consequence of spatial neglect on activities of daily living was evaluated with the Bergego scale (ECB) (Bergego et al. 1995) and the Functional Independence Measure (FIM) (Granger et al. 1986).

**Figure 1 :**
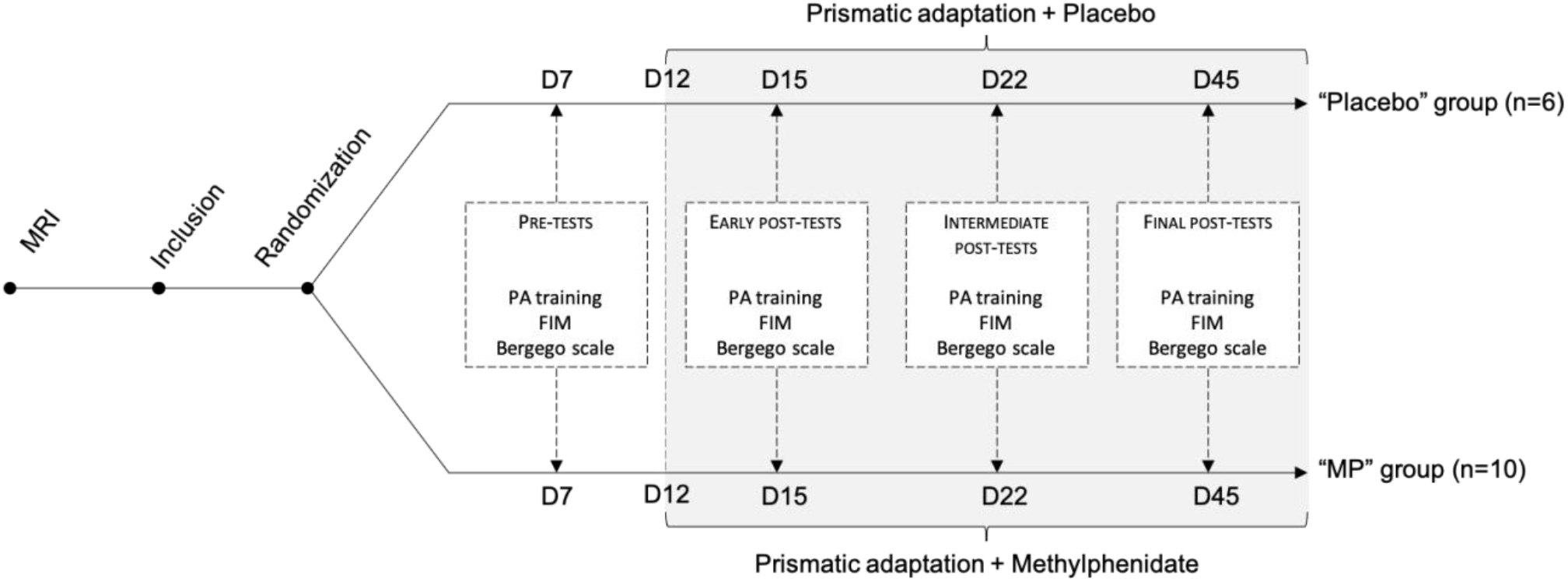
Study design. *FIM: Functional Independence Measure; MP: methylphenidate; PA: prismatic adaptation*. (Adapted from Luauté et al., 2018).

### 2.3 MRI segmentation

All patients performed a 3D T1 MRI. Individual segmentations were created from the MRIs of 16 patients by manually labelling 10 brain structures (anterior caudate nucleus, anterior putamen, ventral, posterior caudate nucleus, posterior putamen) in three dimensions (Figure 2), following a precise delineation protocol (Supplementary material) using ITK-Snap (Yushkevich et al. 2006) (http://www.itksnap.org). The lesion area was also manually segmented. All delineation were performed in native space. The volume of each ROI was extracted. The presence or absence of a lesion within a ROI was dichotomised. A ROI was considered as lesioned if the difference between right and left ROI volume was superior to three time the standard error of the mean of the left ROI volume using the following formula : Volume_left ROI_ – Volume_right ROI_ >3*SEM Volume _left ROI group_.

**Figure 2 :**
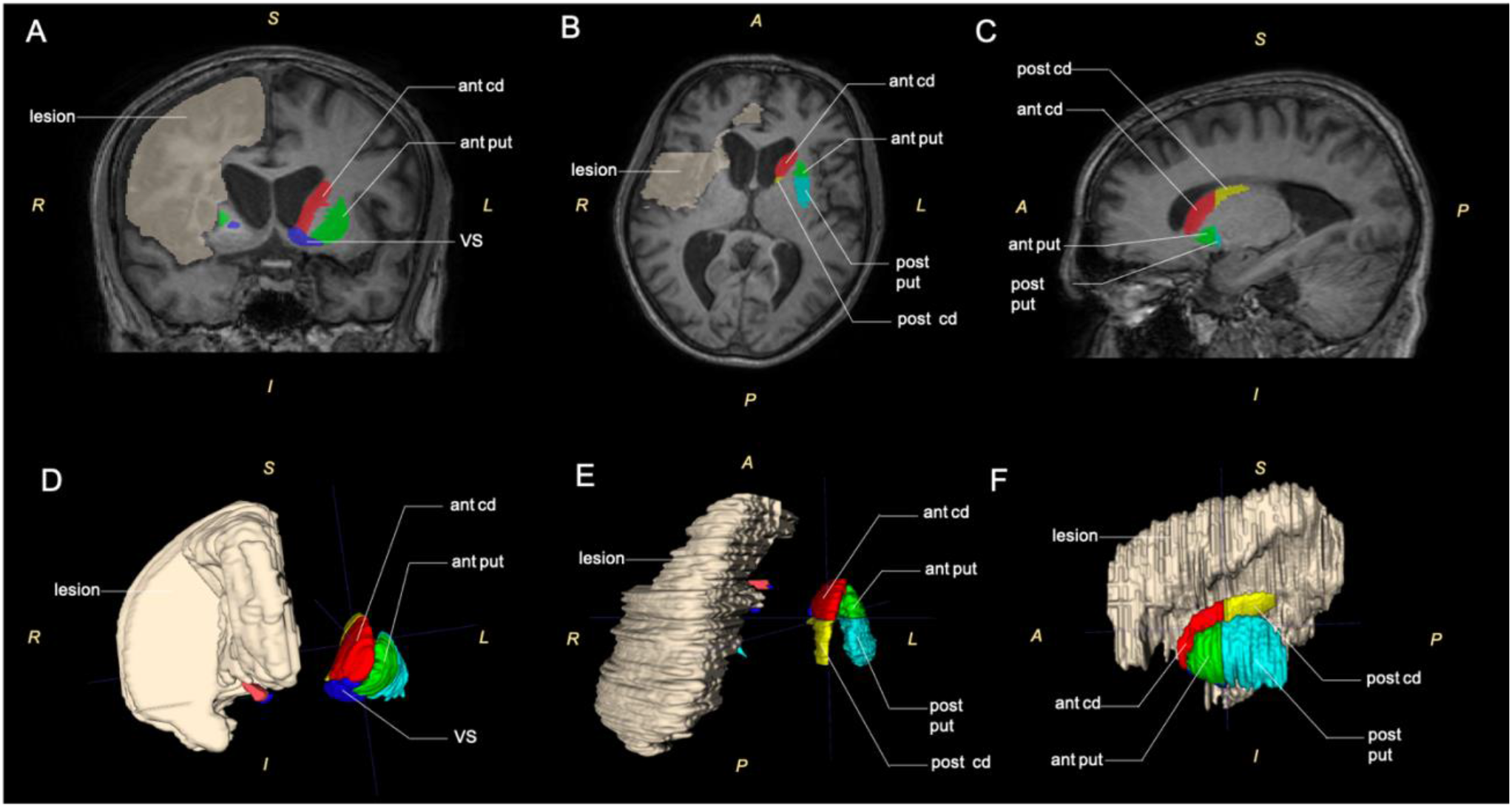
Illustration of an MRI individual segmentation. (A) Coronal, (B) transversal and (C) sagittal MRI section. (D) Anterior, (E) superior and (F) lateral view of 3-dimentional reconstruction of regions of interest. *Ant: anterior; cd: caudate nucleus; put: putamen; VS: ventral striatum*.

### 2.4 Stastistical analysis

All statistical analyses were performed using R software (https://www.r-project.org). The effect of ROI lesion on i) initial severity of spatial neglect, ii) prism adaptation and iii) methylphenidate (MP) efficiency was assessed by non-parametric Mann-Whitney-Wilcoxon test. Clinical scores (ECB, MIF) and the line bisection measure were compared between lesioned and non-lesioned patients (for each ROI). Initial severity of spatial neglect was assessed in the whole cohort at D7 (pre-tests; Figure 1). The effect of striatal lesion on prism adaptation was assessed in the placebo group at D15, D22 and D45. The effect of striatal lesion on MP efficiency was assessed in the MP group at D15, D22 and D45. P-values of less than 0.05 were considered as significant.

## 3 Results

### 3.1 Effect of striatal lesion on initial severity of spatial neglect at baseline (D7)

A significant reduction of mean bisection was observed in patients presenting a lesion in the right ventral striatum (Figure 3A; p<0.05), the right posterior caudate nucleus (Figure 3B; p<0.05) and the right posterior putamen (Figure 3C; p<0.05). There was no significant difference in ECB and FIM.

**Figure 3 :**
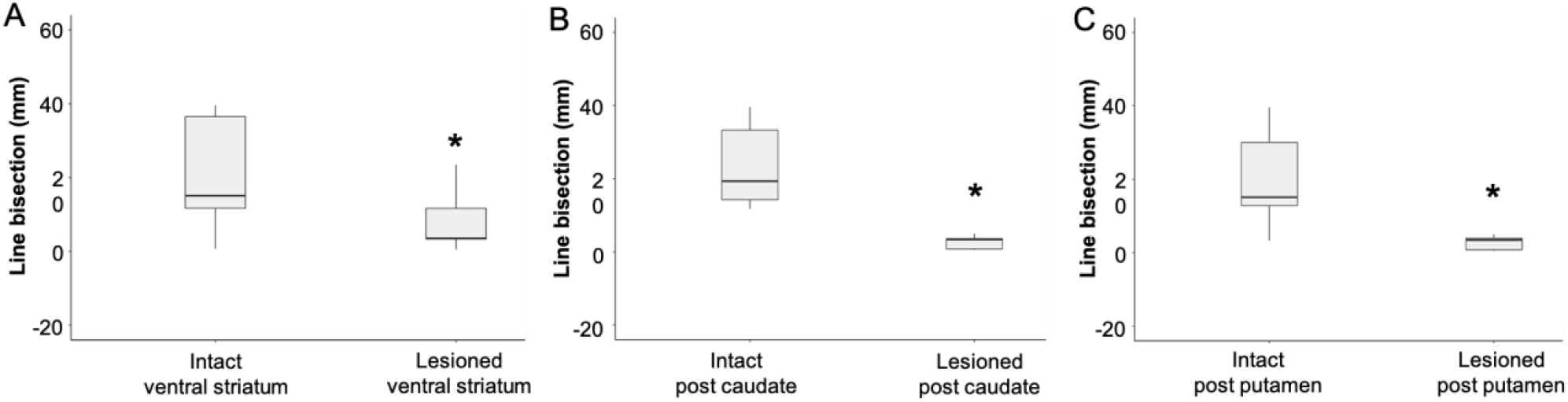
The mean line bisection at D7 was significantly reduced in patients presenting a lesion within (A) the right ventral striatum, (B) the right posterior caudate nucleus and (C) the right posterior putamen. *p<0.05

### 3.2 Effect of striatal lesions on prism adaptation over time

Analysis was conducted in the placebo group (n=6) at D15, D22 and D45. Prism adaptation was evaluated by comparing the variation of absolute value of mean deviation depending on the presence or absence of lesions. There was no significant difference across groups whatever the absence or presence of lesions in any striatal region (i.e. anterior and posterior caudate nucleus, anterior and posterior putamen, ventral striatum) in ECB, FIM and line bisection.

### 3.3 Effect of striatal lesions on MP treatment over time

Analyses were conducted in the MP group (n=10) at D15, D22 and D45.

At D15, a significant reduction of the absolute value of line bisection after MP was observed in patients presenting a lesion in the right ventral striatum (Figure 4A; p<0.05). The variation of absolute value of the mean deviation on the straight-ahead pointing was 4mm in the group with ventral striatal lesion and 14mm in the group without lesion.

**Figure 4 :**
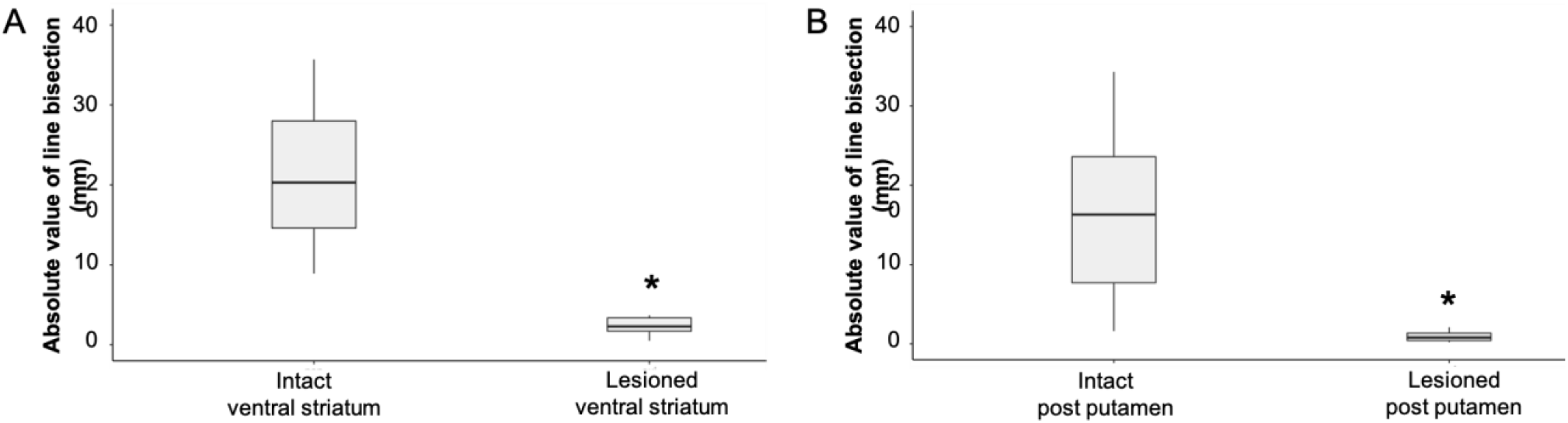
The absolute value of line bisection was significantly reduced in the MP group patients (A) at D15 in patients presenting a lesion within the right ventral striatum and (B) at D45 in patients presenting a lesion within the right posterior putamen. *p<0.05

At D45, a significant reduction of the absolute value of line bisection after MP+PA was observed in patients presenting a lesion in the right posterior putamen (Figure 4B; p<0.05). The variation of absolute value of the mean deviation on the straight-ahead pointing was 1mm in the group with posterior putamen lesion and 20mm in the group without lesion.

## 4 Discussion

This study proposes that the beneficial effect of MP+PA co-administration should require striatal integrity in neglect patients. More specifically, our results suggest that the short-term effect is mediated by the ventral striatum whereas the long-term effect is mediated by the posterior putamen. Patients with severe neglect showed the most benefit from prism adaptation treatment (Gillen et al. 2022), and among then, patients with spared ventral striatal and posterior putamen are most likely to benefit MP+PA co-administration.

### The ventral striatum and the posterior putamen are the substrates for MP+PA action

It is well described now that dopaminergic medication improves motor component of spatial neglect (Geminiani, Bottini, et Sterzi 1998). In addition, PA preferentially improves motor intentional bias than perceptual bias in both healthy and neglect patients (Fortis et al. 2011). Thus, the involvement of the ventral part of the striatum – i.e. an associativo-limbic territory (Alexander, DeLong, et Strick 1986) – during a MP+PA co-administration was surprising.

An *in vivo* PET and pharmacological study in non-human primates has suggested that the beneficial effect of MP could be mediated by the anterior ventral striatum. Indeed, Martinez and colleagues have shown that a systemic administration of MP decreases the fixation of ^11^C-PE2I radiotracer (targeting dopaminergic transporters) within the ventral striatum (Martinez et al. 2020), which is known to be involved in reward processing and reinforcement learning (Tremblay et al. 2015). Altogether, the involvement of the ventral striatum suggests that reinforcement processes could be involved in MP+PA mechanisms. In fact, placing a high reward on certain targets can override intrinsic spatial attention bias (Della Libera et Chelazzi 2006; Kristjánsson et al. 2010; J. Lee et Shomstein 2013; Jeongmi Lee et Shomstein 2014). Reinforcement learning could influence the bottom-up influences of stimulus features on attentional allocation during spatial decision making (Neppi-Mòdona et al. 2020).

Our results also highlighted the role of the posterior putamen in MP+PA beneficial effects, as patients with intact posterior putamen demonstrate a long-term improvement. Interestingly, the MP+PA alleviation seems to concern only the classical tests of spatial neglect (i.e. line bisection) – which are specifically design to detect visuospatial alteration – compared to the different clinical scales – which evaluate the global effect of spatial neglect in daily life). The role of posterior parietal cortex in visuospatial attention has been widely described in the literature (for review Silver, Ress, et Heeger 2005). Interestingly, the posterior putamen receives overlapping projections from parietal areas associated with somatosensory function, resulting in a sensory-motor area (Haber 2003). Thus, the posterior putamen appears to be a key region in the beneficial effect of M+PA in improving visuospatial attention.

### The long-term antero-posterior shift in the striatum during MP+PA co-administration

Our study suggests that the beneficial effect of MP+PA takes place in two stages with the involvement of short-term mechanisms in the ventral striatum (assessed at D15) and long-term mechanisms in the posterior putamen (assessed at D45). Thus, reinforcement processes could be involved at early stage of the MP+PA program and replaced by visuospatial substrates.

This diffuse overlap of corticostriatal projections has been proposed as an explicit substrate for reinforcement learning that directly integrates reward and executive control signals from the dorsolateral prefrontal cortex (dlPFC) (for review Haber et Knutson 2010). There are bilateral convergence zones i) in the ventral striatum receiving projections from OFC and dlPCF, ii) in the putamen receiving projections from dlPCF and parietal cortex (Verstynen 2014; Jarbo et Verstynen 2015). During a new learning, the dlPFC and the striatum (i.e. ventral striatum) are activated. When subjects had to select movements (i.e at the beginning of the MP+PA training until D15), the premotor cortex and mid-putamen are activated. With automatic (overlearned) movements (i.e; at the end of the program at D45), the sensorimoteur and posterior putamen are activated (Jueptner et Weiller 1998). In other words, there is a shift of activation in the anterior-posterior direction within the striatum. This antero-posterior shift within the striatum would explain why an intact ventral striatum could be needed at D15 while an intact posterior putamen seems to be mandatory at D45 for a long-term benefit. Further studies are need to decipher the role of dlPFC in MP+PA beneficial effet in spatial neglect.

## Supporting information

Supplementary : Delineation protocol

## Data Availability

All data produced in the present study are available upon reasonable request to the authors

