## Supplementary : Delineation protocol for "Co-administration of prism adaptation and methylphenidate needs striatal integrity to alleviate spatial neglect"

### Supplementary material: delineation protocol

**Anterior caudate nucleus (left and right)**

| **Orientation of slices** | **Coronal** |
|---|---|
| Anterior border | Most anterior slice when the caudate nucleus is seen |
| Posterior border | Last slice on which the anterior commissure is completely visualized |
| Superior border | White matter |
| Inferior border | From anterior to posterior: white matter, ventral striatum, white matter |
| Medial border | White matter |
| Lateral border | White matter |

**Anterior putamen (left and right)**

| **Orientation of slices** | **Coronal** |
|---|---|
| Anterior border | Most anterior slice when the caudate nucleus is seen |
| Posterior border | Last slice on which the anterior commissure is completely visualized |
| Superior border | White matter |
| Inferior border | White matter |
| Medial border | From anterior to posterior: internal capsule and ventral striatum, pallidum |
| Lateral border | White matter |

**Ventral striatum (left and right)**

The ventral striatum corresponds to the ventro-medial portion of the caudate nucleus and most ventral and medial part of the putamen (Haber, 2003).

| **Orientation of slices** | **Coronal** |
|---|---|
| Anterior border | First slice when the inferior part of the caudate nucleus is visible |
| Posterior border | Last slice on which the anterior commissure is completely visualized |
| Superior border | Anterior striatum (ventral striatum is always inferior to lateral ventricle) |
| Inferior border | From anterior to posterior: white matter, anterior commissure |

| | |
|---|---|
| Medial border | Lateral ventricle, white matter |
| Lateral border | White matter |

**Posterior caudate nucleus (left and right)**

| **Orientation of slices** | **Coronal** |
|---|---|
| Anterior border | Anterior caudate nucleus |
| Posterior border | Last slice when visible |
| Superior border | White matter |
| Inferior border | From anterior to posterior: white matter, thalamus, white matter |
| Medial border | Lateral ventricle |
| Lateral border | White matter |

**Posterior putamen**

| **Orientation of slices** | **Coronal** |
|---|---|
| Anterior border | Anterior putamen |
| Posterior border | Last slice when visible |
| Superior border | White matter |
| Inferior border | White matter |
| Medial border | From anterior to posterior: white matter, pallidum, white matter |
| Lateral border | White matter |
